# Student Scholarly Research Programs in US Medical Schools: Cross-sectional Web Audit

**DOI:** 10.64898/2026.03.03.26347497

**Authors:** Dongyoon Lee, Changyoon Lee, Sarah Soyeon Oh, Keeheon Lee, Chul S Hyun, Jae Il Shin, Shinki An, John PA Ioannidis

## Abstract

**Background:** Participating in research during medical school is supported by institutional programs and may influence subsequent professional development.

**Objective:** We aimed to describe the current status and heterogeneity of scholarly research programs for medical students in the United States, including expectations, support, and key structural features.

**Methods:** We conducted a cross-sectional web audit of official webpages for all accredited US MD- and DO-granting medical schools (search performed September 2024 to January 2025). Extracted variables included participation requirements, mentorship, timing and duration (overall and dedicated research time), expected scholarly outputs, funding sources, stipend information, and stated program goals. We compared Carnegie tier R1 (Very high research activity) versus other institutions, QS Top-50 versus other institutions, and MD versus DO schools using χ^2^/Fisher exact tests for 2×2 tables and exact trend or Freeman–Halton tests for multicategory variables.

**Results:** Programs were identified for all 202 institutions. Funding was explicitly mentioned by 61.9% (125/202) of programs, 27.0% (51/189) were compulsory, 98.9% (188/190) reported faculty mentorship, and 91.0% (171/188) were exclusive for medical students. Program duration, dedicated time, expected outcomes, stipend reporting, funding sources, and stated goals varied widely. Carnegie R1 institutions had longer duration (P=.002) and tended to report external funding more often than other institutions (25/104, 24.0% vs 9/98, 9.2%; OR 3.13, 95% CI 1.38-7.10; P=.008). QS Top-50 institutions were more likely to require compulsory participation than other institutions (11/19, 57.9% vs 40/170, 23.5%; OR 4.47, 95% CI 1.68-11.87; P=.003). No significant differences were observed between MD and DO programs across most measured characteristics.

**Conclusions:** Scholarly research programs for medical students are ubiquitous across US medical schools but heterogeneous in structure, expectations, and support. Research-intensive and top-ranked institutions may have more external funding and sometimes may put together longer and compulsory programs Further evaluation of student experiences and outcomes is warranted.

## Introduction

Engaging in scholarly research during medical school may help inspire and prepare some students to pursue a physician-scientist career [1], but may also be beneficial for physicians who will go to health care without research aspirations. The research experience may translate into improved patient care if it can foster the ability to integrate science-based, evidence-based reasoning into the clinical decision-making process [2,3]. Early exposure to research may cultivate a deeper understanding of the scientific method, enhance problem-solving abilities, and encourage a lifelong commitment to learning and discovery within the medical profession [4,5]. The physician-scientist pipeline has long been under threat [6], but it is unclear which measures might help sustain it and whether engaging students in research during medical school is an effective measure in this regard. For the average medical student, the outcomes are even more uncertain. A review of 39 studies on ‘scholarly concentration’ programs found mixed results [7]. Some studies suggested that some students did go into academic careers, publish papers or made research presentations, or were helped in their clinical specialty choice [7]. However, other studies noted the burden of mixing such programs with the already high demands of clinical training and the effort required to complete scholarly projects [7].

The literature also highlights substantial heterogeneity in program designs [4]. For example, an evaluation of programs in 15 different US medical schools showed diversity in program structures, focus, duration, centralization, capstone requirement, faculty involvement, outcome definitions, and evaluation rigor [8]. At the curricular level, schools have adopted diverse approaches ranging from elective summer programs, multi-year mentored projects, and scholarly concentration tracks. Each has different resource requirements and potential equity implications (i.e., reliance on external funding, variable stipend support, and differences in protected time) [8,9].

As these programs have become practically ubiquitous across medical schools in the USA, it is important to gain a contemporary, national-level characterization of what medical schools publicly report about their student scholarly research programs and how consistently key program features are described across institutions. Prior work has underscored both the importance of scholarship opportunities and the difficulty of drawing cross-institutional conclusions when program descriptors are inconsistent or incomplete [7]. A systematic assessment of publicly available information can therefore serve as a pragmatic benchmarking tool and a first step toward identifying remediable gaps in transparency and standardization. It may also help to perform comparisons across different types of institutions in terms of key features of the programs that they offer.

In this cross-sectional web audit, we aimed to characterize the publicly disclosed structural features of medical student scholarly research programs across all US medical schools. We focus on program identification and coverage as well as key descriptors such as funding or stipend reporting, duration and timing, mentorship, expected outcomes, and stated goals. We further investigated whether there were differences in reported program characteristics based on institutional research intensity, global ranking strata, and type of degree.

## Methods

### Study Design

We conducted a cross-sectional web audit of publicly available, official institutional webpages. All US medical schools accredited to grant MD or DO degrees at the time the web audit was performed (September 2024 - January 2025) were included. The school list was acquired from the Liaison Committee on Medical Education (LCME) and the American Association of Colleges of Osteopathic Medicine (AACOM), and each institution’s active status was verified during extraction. We defined a ‘scholarly research program’ as an institutionally described, structured pathway intended for students that includes explicit expectations (i.e., duration or milestones) and institutional supports (i.e., mentorship, protected time, or funding). Ad hoc research opportunities without a defined program structure were not included. We report methods and results in alignment with STROBE principles for cross-sectional studies (Multimedia Appendix 2). We did not pre-register a protocol, as we came across peculiarities in specific websites and tried to capture maximal information.

### Search Strategy and Data Extraction

For each institution, we first navigated from the official medical school homepage to pages describing research or scholarly requirements. If not located via navigation, we performed site-specific searches using standardized keywords (‘scholarly concentration’, ‘research track’, ‘summer research’, ‘student research’, ‘student research program’, ‘medical student research’, and ‘medical student research program’). When multiple programs were listed for a school, we created a single school-level record using the earliest start year, latest end year, and the maximum duration (overall and dedicated). Binary features were coded as present if offered by any program (e.g., compulsory if any program was compulsory; stipend available if any program offered a stipend), since short programs were usually brief components included within longer or extended pathways and were often not distinct programs in practice.

### Data Extraction

A standardized extraction sheet was developed a priori. Two reviewers (DL and CL) piloted the form on an initial subset of schools, refined variable definitions, and then extracted data in duplicate for a prespecified subset to assess consistency. Discrepancies were resolved by consensus, with discussion with a third investigator (JS) when needed. Extracted variables included whether student participation was compulsory, whether the program included summer projects, the presence of faculty mentors, program start and end years, overall duration, dedicated research time, expected scholarly outputs, funding sources, stipend amount, and stated program goals.

### Statistical Analysis

We compared program characteristics by institutional research intensity (Carnegie tier R1 [very high research activity] vs others) [19], global ranking status (QS ranking in the Top-50 vs others) [20], and degree type (MD vs DO). For 2×2 tables, we used χ^2^ tests when all expected cell counts were ≥5; otherwise, we used Fisher exact tests. For ordered categorical variables, we used two-sided Cochran-Armitage exact trend tests; for nominal multicategory variables, we used two-sided exact Freeman-Halton tests. For ordered variables with a significant trend, we estimated effect sizes using proportional odds models, which yield a single common odds ratio summarizing the odds of being in a higher category. When cutpoint-specific ORs from separate binary logistic regressions at each cumulative threshold showed substantial heterogeneity (indicating violation of the proportional odds assumption), we report the cutpoint-specific ORs instead. Analyses were conducted using R (version 4.4.2). We report absolute counts and percentages with variable-specific denominators. Because comparisons were hypothesis-generating across multiple program features, we emphasize effect sizes and consistency of patterns rather than P values alone. P-values of less than .005 are considered fully statistically significant and P-values of .05 to .005 are considered to have suggestive statistical significance [21].

### Ethical Considerations

This study used only publicly available information from official medical school program webpages and did not involve human participants, patient data, or interaction with individuals. Therefore, institutional review board approval and informed consent were not required.

## Results

### Webpage identification/coverage

We conducted a comprehensive review of official webpages describing medical student scholarly research programs for all 202 accredited US MD and DO-granting medical schools. Qualifying programs were identified for 202/202 institutions (100%). For each institution, the primary program webpage was identified, and data were extracted from the most current page available during the review period (September 2024 - January 2025). For 14 schools that provided multiple programs, we abstracted all the programs provided.

Institutions varied in how extensively they documented program details, program attributes were frequently not documented on webpages, leading to variable-specific denominators across measures (i.e., compulsory participation reported for 189/202 schools, mentorship for 190/202, and exclusivity for 188/202). To characterize documentation completeness more explicitly, we summarized missingness by variable and compared missingness patterns across institutional subgroups (Multimedia Appendix 1). Differences in missingness between institutional subgroups reached P<.005 for starting year (R1: 14/104 vs non-R1: 33/98, P=.001; MD: 24/158 vs DO: 23/44, P<.001), ending year (MD: 29/158 vs DO: 23/44,

P<.001), and duration (MD: 16/158 vs DO: 14/44, P<.001) (Multimedia Appendix 1).

### Program characteristics: overall

As shown in Table 1, programs in half of the schools mentioned a program director, nearly two-thirds explicitly mentioned funding, and slightly over one-quarter were compulsory.

**Table 1.**
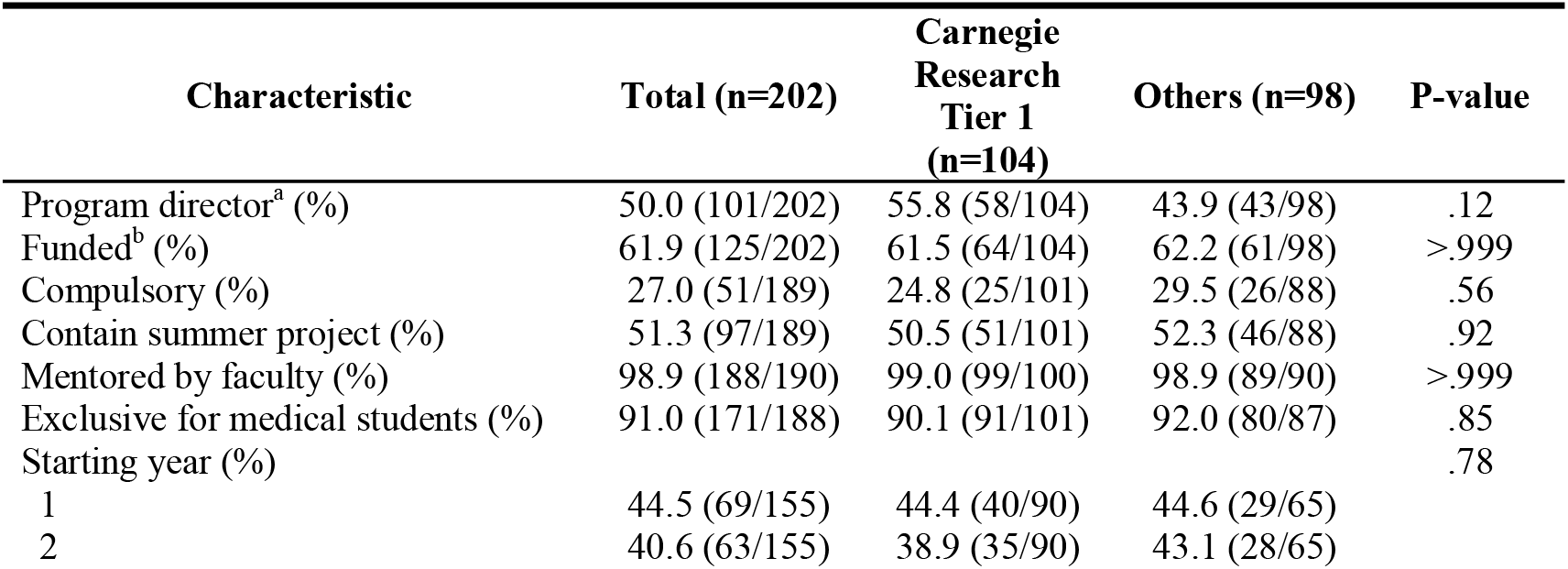
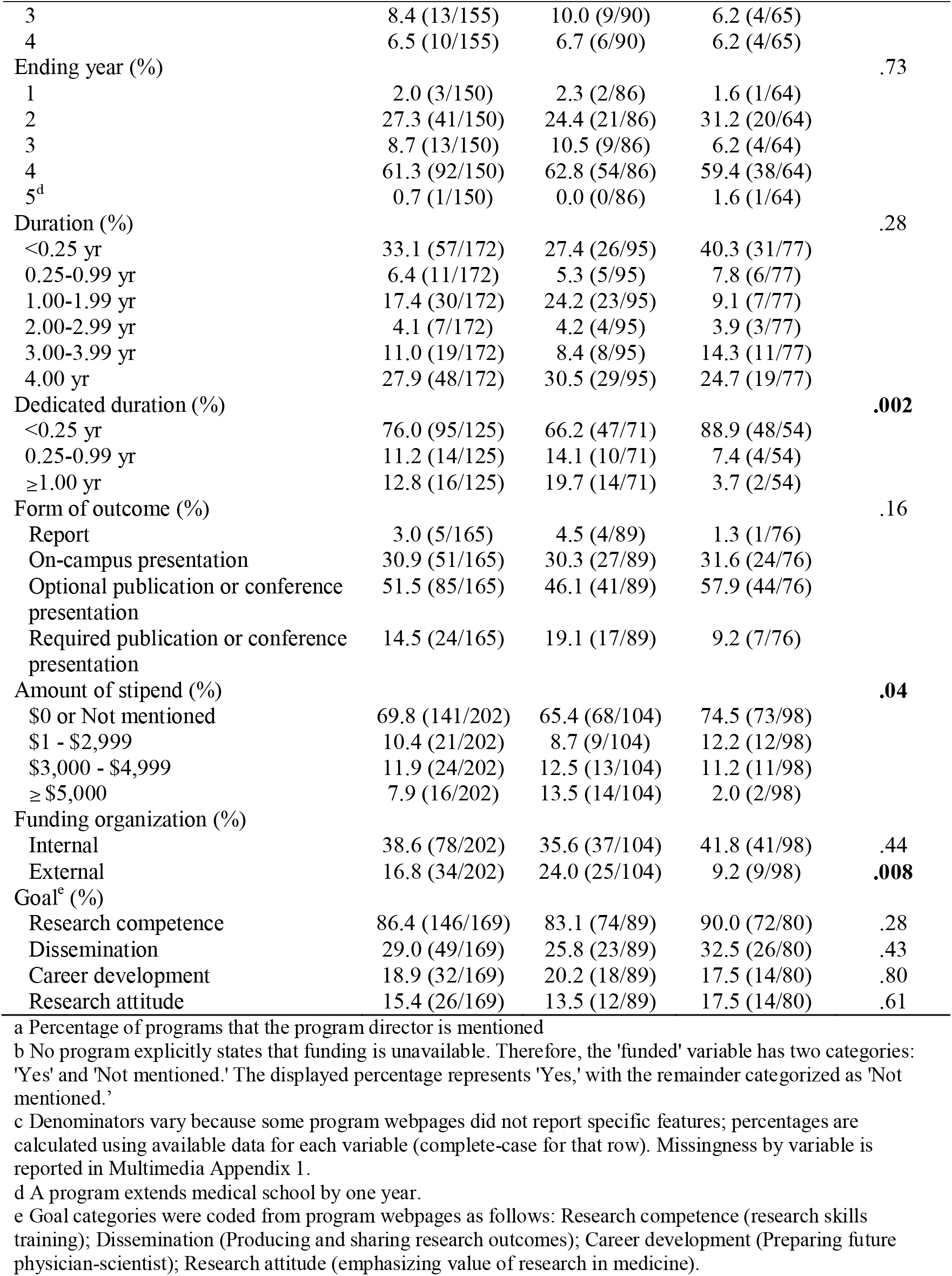
Program characteristics: overall and by Carnegie tier for their research concentration.

Almost all programs explicitly described faculty mentorship and were exclusive to medical students. About half included a summer project component.

Structure and time commitment varied substantially. Overall program duration ranged from short-term experiences of less than 0.25 year in a third of the programs to longitudinal programs spanning 4.0 years or more in almost another third. In contrast, dedicated research time was most commonly (in over three-quarters) less than 0.25 year. Expected scholarly outputs also differed across programs. Among described outcomes (n=165), slightly over half expected publication or conference presentation as optional (51.5%), while one out of seven required a publication or conference presentation (14.5%). Other programs emphasized on-campus presentations (30.9%) or written reports (3.0%).

Financial support was inconsistently reported. Stipend was frequently absent or reported as $0 in more than two-thirds of the programs. Among programs reporting stipends, amounts ranged substantially, with 16 programs offering $5,000 or more. Funding sources were also heterogeneous, with internal funding noted in 38.6% and external funding in 16.8%.

### Program characteristics in institutional subgroups

As shown in Table 1, Carnegie tier R1 institutions more frequently reported longer dedicated research duration (OR 4.23, 95% CI 1.59–11.24; P=.004). Moreover, there was a suggestion that programs at Carnegie tier R1 institutions reported external funding more often than others (25/104, 24.0% vs 9/98; OR 3.13, 95% CI 1.38-7.10; P=.008), and higher stipends (P=.04) (Table 1). As cutpoint-specific odds ratios varied substantially across the stipend distribution (thus, the proportional odds assumption was not met), we report these separately:

R1 institutions more often tended to report stipends of ≥ $5,000 (OR 7.47, 95% CI 1.65-33.77; P=.009) and ≥$3,000 (OR 2.29, 95% CI 1.10-4.76; P=.03), but the odds of reporting any versus no stipend didn’t differ significantly (OR 1.55, 95% CI 0.84-2.84; P=.16).

As shown in Table 2, there were only 19 institutions that were ranked among the top-50 worldwide according to the QS ranking, thus comparisons against other institutions may be tenuous due to limited precision. Programs at QS Top-50 institutions were more likely to be compulsory than those at others (11/19, 57.9% vs 40/170, 23.5%; OR 4.47, 95% CI 1.68-11.87; P=.003). They also tended to offer longer dedicated research duration (≥1.00 yr 4/16, 25.0% vs 12/109, 11.0%; OR 4.32, 95% CI 1.58–11.83; P=.004), more likely to start in a later year (OR 2.69, 95% CI 1.06-6.84; P=.04) (Table 2), were less likely to mention a stipend (P=.03), and more likely to start in a later year (OR 2.69, 95% CI 1.06-6.84; P=.04).

**Table 2.** Program characteristics by QS Top-50 status.

| Characteristic | QS Top 50<br>(n=19) | Others<br>(n=183) | P-value |
| --- | --- | --- | --- |
| Program director<sup>a</sup> (%) | 47.4 (9/19) | 50.3 (92/183) | >.999 |
| Funded<sup>b</sup> (%) | 42.1 (8/19) | 63.9 (117/183) | .11 |
| Compulsory (%) | 57.9 (11/19) | 23.5 (40/170) | **.003** |
| Contain summer project (%) | 36.8 (7/19) | 52.9 (90/170) | .28 |
| Mentored by faculty (%) | 100.0 (19/19) | 98.8 (169/171) | >.999 |
| Exclusive for medical students (%) | 94.7 (18/19) | 90.5 (153/169) | >.999 |
| Starting year (%) |  |  | **.04** |
| 1 | 27.8 (5/18) | 46.7 (64/137) |  |
| 2 | 38.9 (7/18) | 40.9 (56/137) |  |
| 3 | 22.2 (4/18) | 6.6 (9/137) |  |
| 4 | 11.1 (2/18) | 5.8 (8/137) |  |
| Ending year (%) |  |  | .78 |
| 1 | 0.0 (0/16) | 2.2 (3/134) |  |
| 2 | 25.0 (4/16) | 27.6 (37/134) |  |
| 3 | 25.0 (4/16) | 6.7 (9/134) |  |
| 4 | 50.0 (8/16) | 62.7 (84/134) |  |
| 5<sup>d</sup> | 0.0 (0/16) | 0.7 (1/134) |  |
| Duration (%) |  |  | .45 |
| <0.25 yr | 11.8 (2/17) | 35.5 (55/155) |  |
| 0.25-0.99 yr | 23.5 (4/17) | 4.5 (7/155) |  |
| 1.00-1.99 yr | 41.2 (7/17) | 14.8 (23/155) |  |
| 2.00-2.99 yr | 5.9 (1/17) | 3.9 (6/155) |  |
| 3.00-3.99 yr | 11.8 (2/17) | 11.0 (17/155) |  |
| 4.00 yr | 5.9 (1/17) | 30.3 (47/155) |  |
| Dedicated duration (%) |  |  | **.01** |
| <0.25 yr | 43.8 (7/16) | 80.7 (88/109) |  |
| 0.25-0.99 yr | 31.2 (5/16) | 8.3 (9/109) |  |
| ≥1.00 yr | 25.0 (4/16) | 11.0 (12/109) |  |
| Form of outcome (%) |  |  | .46 |
| Report | 6.2 (1/16) | 2.7 (4/149) |  |
| On-campus presentation | 43.8 (7/16) | 29.5 (44/149) |  |
| Optional publication or conference presentation | 43.8 (7/16) | 52.3 (78/149) |  |
| Required publication or conference presentation | 6.2 (1/16) | 15.4 (23/149) |  |
| Amount of stipend (%) |  |  | **.03** |
| \$0 or Not mentioned | 94.7 (18/19) | 67.2 (123/183) | |
| \$1 - \$2,999 | 0.0 (0/19) | 11.5 (21/183) | |
| \$3,000 - \$4,999 | 0.0 (0/19) | 13.1 (24/183) | |
| ≥ \$5,000 | 5.3 (1/19) | 8.2 (15/183) | |
| Funding organization (%) |  |  |  |
| Internal | 31.6 (6/19) | 39.3 (72/183) | .68 |
| External | 21.1 (4/19) | 16.4 (30/183) | .53 |
| Goal<sup>e</sup> (%) |  |  |  |
| Research competence | 76.5 (13/17) | 87.5 (133/152) | .26 |
| Dissemination | 35.3 (6/17) | 28.3 (43/152) | .58 |
| Career development | 23.5 (4/17) | 18.4 (28/152) | .53 |
| Research attitude | 5.9 (1/17) | 16.4 (25/152) | .48 |

a Percentage of programs that the program director is mentioned
b No program explicitly states that funding is unavailable. Therefore, the 'funded' variable has two categories: 'Yes' and 'Not mentioned.' The displayed percentage represents 'Yes,' with the remainder categorized as 'Not mentioned.'
c Denominators vary because some program webpages did not report specific features; percentages are calculated using available data for each variable (complete-case for that row). Missingness by variable is reported in Multimedia Appendix 1.
d A program extends medical school by one year.
e Goal categories were coded from program webpages as follows: Research competence (research skills training); Dissemination (Producing and sharing research outcomes); Career development (Preparing future physician-scientist); Research attitude (emphasizing value of research in medicine).

In contrast, comparisons between MD vs DO programs showed no statistically significant or suggestive differences across the measured characteristics (Table 3).

**Table 3.** Program characteristics by degree type.

| Characteristic | Doctor of<br>Medicine<br>(n=158) | Doctor of<br>Osteopathic<br>Medicine<br>(n=44) | P-value |
| --- | --- | --- | --- |
| Program director<sup>a</sup> (%) | 53.8 (85/158) | 36.4 (16/44) | .06 |
| Funded<sup>b</sup> (%) | 61.4 (97/158) | 63.6 (28/44) | .92 |
| Compulsory (%) | 27.2 (41/151) | 26.3 (10/38) | >.999 |
| Contain summer project (%) | 52.3 (79/151) | 47.4 (18/38) | .72 |
| Mentored by faculty (%) | 98.7 (148/150) | 100.0 (40/40) | >.999 |
| Exclusive for medical students (%) | 91.4 (138/151) | 89.2 (33/37) | .75 |
| Starting year (%) |  |  | .79 |
| 1 | 44.0 (59/134) | 47.6 (10/21) |  |
| 2 | 41.8 (56/134) | 33.3 (7/21) |  |
| 3 | 6.7 (9/134) | 19.0 (4/21) |  |
| 4 | 7.5 (10/134) | 0.0 (0/21) |  |
| Ending year (%) |  |  | .81 |
| 1 | 2.3 (3/129) | 0.0 (0/21) |  |
| 2 | 27.1 (35/129) | 28.6 (6/21) |  |
| 3 | 8.5 (11/129) | 9.5 (2/21) |  |
| 4 | 62.0 (80/129) | 57.1 (12/21) |  |
| 5<sup>d</sup> | 0.0 (0/129) | 4.8 (1/21) |  |
| Duration (%) |  |  | .15 |
| <0.25 yr | 31.0 (44/142) | 43.3 (13/30) |  |
| 0.25-0.99 yr | 6.3 (9/142) | 6.7 (2/30) |  |
| 1.00-1.99 yr | 17.6 (25/142) | 16.7 (5/30) |  |
| 2.00-2.99 yr | 4.2 (6/142) | 3.3 (1/30) |  |
| 3.00-3.99 yr | 10.6 (15/142) | 13.3 (4/30) |  |
| 4.00 yr | 30.3 (43/142) | 16.7 (5/30) |  |
| Dedicated duration (%) |  |  | .87 |
| <0.25 yr | 75.0 (78/104) | 81.0 (17/21) |  |
| 0.25-0.99 yr | 12.5 (13/104) | 4.8 (1/21) |  |
| ≥1.00 yr | 12.5 (13/104) | 14.3 (3/21) |  |
| Form of outcome (%) |  |  | .09 |
| Report | 3.1 (4/129) | 2.8 (1/36) |  |
| On-campus presentation | 33.3 (43/129) | 22.2 (8/36) |  |
| Optional publication or conference presentation | 46.5 (60/129) | 69.4 (25/36) |  |
| Required publication or conference presentation | 17.1 (22/129) | 5.6 (2/36) |  |
| Amount of stipend (%) |  |  | .08 |
| \$0 or Not mentioned | 67.1 (106/158) | 79.5 (35/44) | |
| \$1 - \$2,999 | 10.8 (17/158) | 9.1 (4/44) | |
| \$3,000 - \$4,999 | 12.7 (20/158) | 9.1 (4/44) | |
| ≥ \$5,000 | 9.5 (15/158) | 2.3 (1/44) | |
| Funding organization (%) |  |  |  |
| Internal | 36.7 (58/158) | 45.5 (20/44) | .38 |
| External | 19.6 (31/158) | 6.8 (3/44) | .07 |
| Goal<sup>e</sup> (%) |  |  |  |
| Research competence | 84.3 (113/134) | 94.3 (33/35) | .17 |
| Dissemination | 26.9 (36/134) | 37.1 (13/35) | .33 |
| Career development | 19.4 (26/134) | 17.1 (6/35) | .95 |
| Research attitude | 14.9 (20/134) | 17.1 (6/35) | .95 |

a Percentage of programs that the program director is mentioned
b No program explicitly states that funding is unavailable. Therefore, the 'funded' variable has two categories: 'Yes' and 'Not mentioned.' The displayed percentage represents 'Yes,' with the remainder categorized as 'Not mentioned.'
c Denominators vary because some program webpages did not report specific features; percentages are calculated using available data for each variable (complete-case for that row). Missingness by variable is reported in Multimedia Appendix 1.
d A program extends medical school by one year.
e Goal categories were coded from program webpages as follows: Research competence (research skills training); Dissemination (Producing and sharing research outcomes); Career development (Preparing future physician-scientist); Research attitude (emphasizing value of research in medicine)

## Discussion

### Principal Findings and Institutional Differences

In this cross-sectional web audit of scholarly research programs across all 202 US MD- and DO-granting medical schools, formal programs were publicly identifiable at every institution. Core elements of programs were commonly documented. Most programs explicitly described faculty mentorship and were exclusive to medical students, and funding was mentioned on the majority of program webpages. However, beyond shared features, programs showed heterogeneity in structure, including duration and dedicated research time, expected scholarly outputs, and reporting of stipends and funding sources.

We also observed some notable subgroup differences by institutional context. Research-intensive institutions (Carnegie tier R1) tended to report external funding more frequently and to have higher stipend amounts, and they had long dedicated durations more frequently than non-R1 institutions. Top-ranked institutions (QS Top-50) were more likely to require compulsory participation and also tended to have longer dedicated durations. These patterns are plausibly explained by differences in research infrastructure, availability of external grant mechanisms, and institutional priorities [4,10]. For example, R1 institutions may have more established pathways for connecting students to externally supported research environments, and highly ranked institutions may integrate scholarship more tightly into curricula and graduation expectations [10–12]. Suggestive differences in research stipends need to be seen with caution, as this information was missing for many programs and only a few institutions are among the QS Top-50.

The observed heterogeneity also likely reflects genuine differences in curricular design and mission. How schools choose to balance research training with clinical and foundational science requirements largely affects the program design [13]. For example, research on medical education in Asia points to how factors like cultural values, leadership gaps, and differing institutional missions can shape what schools choose to emphasize, often resulting in less focus on research in favor of clinical training or policy-driven goals [14].

Importantly, our findings highlight variation in documentation practice. Some program components critical to student decision-making, such as stipend availability, the source of funding, and whether dissemination is required, were frequently not explicitly stated on webpages. This matters because program webpages often serve as the first point of contact for students and applicants, shaping expectations about time commitment, mentorship access, and the feasibility of participation (particularly for students with financial constraints). In prior recruitment contexts, applicants report using program websites during key decision points (i.e., where to apply and interview), and many perceive websites as incomplete or missing desired information [15–17]. Because financial limitations and lack of protected time are consistently identified as barriers to student research participation, omission of stipend and funding details may disproportionately deter students with fewer financial resources from pursuing these opportunities [4,18].

### Aligning standards, practices, and goals with feasible training

Student research programs warrant careful alignment between stated goals, required activities, and the time and infrastructure available. For example, requirements to publish or present at conferences may be unrealistic when protected research time is brief. Even worse, publication-oriented expectations may inadvertently incentivize superficial projects or poor-quality research practices. This is a concern not only for medical students but even for later stage medical trainees, e.g. residents, where mandates for publications may generate mostly small, unreliable, poorly done studies and equally bad publications [22]. A scoping review shows that medical students are unaware of the problems with predatory publishing and some had already used predatory journals or were thinking of using them [23].

Eventually, while many programs articulate broad “research competence” goals, meaningful attainment may be unlikely without longitudinal mentorship, structured training, and adequate protected time. How students should be exposed to scholarship also remains contested: in some settings, it may be more educationally efficient to invest the limited available time and resources towards improving research literacy and critical appraisal skills rather than requiring students to conduct original research projects themselves.

### Transparent minimum reporting set for student-facing webpages

Because program webpages function as student-facing commitments, a standardized minimum reporting set could improve transparency and expectation-setting and reduce mismatches between perceived and actual program demands. At a minimum, consistent reporting of eligibility requirements (including compulsory features), protected time (if any, and if so, when and how), mentorship structure (e.g. mentor assignment approach, expected meeting frequency, any availability of methods/statistical support), expected outputs, and funding availability would better support informed participation. Transparent disclosure may also be important for equity, as unclear stipend and funding information can disproportionately constrain students with limited financial flexibility.

### Limitations

This study has several limitations inherent to a web audit design. First, our findings depend on publicly available, self-presented webpage information, which may be incomplete, outdated, or selectively emphasized, and thus may not reflect actual program implementation. Second, missingness was substantial for some features and some observed differences between different institutions could reflect documentation practices rather than true program differences. Finally, we did not assess program quality, student experiences, mentorship effectiveness, or downstream outcomes (i.e., research competence, publications, residency placement, or long-term academic career trajectories), which cannot be inferred reliably from webpages alone.

### Future Directions

Future research should link program structures to outcomes and experiences. Combining surveys or interviews of program leaders and students, structured evaluation of mentorship and protected time, and objective outcomes such as dissemination or research skill assessments could clarify which program elements are most strongly associated with educational benefit, if any, and which might lead to waste of resources and waste of precious time for already overburdened medical students. Additionally, the development and validation of standardized measures for student research experience and competency could enable cross-institutional comparisons while respecting differing institutional missions.

## Conclusions

Student scholarly research programs are ubiquitous across US medical schools but vary widely in publicly stated structure, expectations, and support. Research-intensive institutions may more often report external funding and tend to be longer, and top-ranked institutions more often mandate participation. More transparent and standardized reporting of key program features may improve expectation-setting and access to research opportunities.

Further evaluations incorporating student experiences and outcomes are warranted.

## Supporting information

Multimedia Appendix 1

Multimedia Appendix 2

Multimedia Appendix 3

## Abbreviations

DO: Doctor of Osteopathic Medicine
MD: Doctor of Medicine
QS: Quacquarelli Symonds (QS World University Rankings)
R1: Carnegie Classification “Very High Research Activity”

## Author’s Contributions

DL and CL contributed equally to this work and share co–first authorship. DL and CL contributed to investigation and data curation (search and data extraction), performed the formal analysis, and drafted the manuscript (writing - original draft). JS contributed to conceptualization, methodology, validation (data extraction consensus), supervision, and project administration, and provided critical manuscript review and editing (writing - review & editing). JPI contributed to methodology, supervision and oversight of formal analysis and participated in manuscript revision (writing - review & editing). SA, SSO, KL, and CSH contributed to methodology and provided feedback and manuscript revision (writing - review & editing). All authors reviewed and approved the final manuscript for submission.

## Data Availability

The raw data set is available from the multimedia appendix 3.

## References

1. Solomon SS, Tom SC, Pichert J, Wasserman D, Powers AC. Impact of medical student research in the development of physician-scientists. J Investig Med 2003 May;51(3):149–156.

2. Sackett DL, Rosenberg WMC, Gray JAM, Haynes RB, Richardson WS. Evidence based medicine: what it is and what it isn’t. BMJ British Medical Journal Publishing Group; 1996 Jan 13;312(7023):71–72.

3. Montori VM, Guyatt GH. Progress in Evidence-Based Medicine. JAMA 2008 Oct 15;300(15):1814–1816.

4. Chang Y, Ramnanan CJ. A Review of Literature on Medical Students and Scholarly Research: Experiences, Attitudes, and Outcomes. Acad Med 2015 Aug;90(8):1162.

5. Norman GR, Shannon SI. Effectiveness of instruction in critical appraisal (evidence-based medicine) skills: a critical appraisal. CMAJ Can Med Assoc J 1998 Jan 27;158(2):177–181.

6. Milewicz DM, Lorenz RG, Dermody TS, Brass LF. Rescuing the physician-scientist workforce: the time for action is now. J Clin Invest 2015 Oct 1;125(10):3742–3747.

7. Bierer SB, Chen HC. How to Measure Success: The Impact of Scholarly Concentrations on Students—A Literature Review. Acad Med 2010 Mar;85(3):438.

8. Green EP, Borkan JM, Pross SH, Adler SR, Nothnagle M, Parsonnet J, Gruppuso PA. Encouraging scholarship: medical school programs to promote student inquiry beyond the traditional medical curriculum. Acad Med 2010 Mar;85(3):409–418.

9. Boninger M, Troen P, Green E, Borkan J, Lance-Jones C, Humphrey A, Gruppuso P, Kant P, McGee J, Willochell M, Schor N, Kanter SL, Levine AS. Implementation of a longitudinal mentored scholarly project: an approach at two medical schools. Acad Med 2010 Mar;85(3):429–437.

10. Nguyen M, Chaudhry SI, Asabor E, Desai MM, Lett E, Cavazos JE, Mason HRC, Boatright D. Variation in Research Experiences and Publications During Medical School by Sex and Race and Ethnicity. JAMA Netw Open 2022 Oct 25;5(10):e2238520.

11. Lauer MS, Roychowdhury D. Inequalities in the distribution of National Institutes of Health research project grant funding. Isales C, Zaidi M, Peifer M, editors. eLife 2021 Sep 3;10:e71712.

12. Jalali MS, Hasgul Z. Potential Trade-Offs of Proposed Cuts to the US National Institutes of Health. JAMA Health Forum 2025 Jul 25;6(7):e252228.

13. Harden RM, Sowden S, Dunn WR. Educational strategies in curriculum development: the SPICES model. Med Educ 1984 Jul;18(4):284–297.

14. Majumder MA. Issues and priorities of medical education research in Asia. Ann Acad Med Singap. 2004;33(2):257–263.

15. Embi PJ, Desai S, Cooney TG. Use and Utility of Web-Based Residency Program Information: A Survey of Residency Applicants. J Med Internet Res 2003 Sep 25;5(3):e22.

16. Gaeta TJ, Birkhahn RH, Lamont D, Banga N, Bove JJ. Aspects of residency programs’ web sites important to student applicants. Acad Emerg Med Off J Soc Acad Emerg Med 2005 Jan;12(1):89–92.

17. Smith BB, Long TR, Tooley AA, Doherty JA, Billings HA, Dozois EJ. Impact of Doximity Residency Navigator on Graduate Medical Education Recruitment. Mayo Clin Proc Innov Qual Outcomes 2018 Jun 1;2(2):113–118.

18. Mayne C, Bates H, Desai D, Martin P. A Review of the Enablers and Barriers of Medical Student Participation in Research. Med Sci Educ 2024 Dec 1;34(6):1629–1639.

19. American Council on Education; Carnegie Foundation for the Advancement of Teaching. Institution Search. Carnegie Classification of Institutions of Higher Education. Available from: https://carnegieclassifications.acenet.edu/institutions/?inst=&research2025%5B%5D=1 [accessed Jan 7, 2025]

20. Quacquarelli Symonds (QS). QS World University Rankings 2026: Top global universities. TopUniversities. Published on: 19 June 2025. Available from: https://www.topuniversities.com/world-university-rankings?tab=indicators&sort_by=rank&order_by=asc [accessed Aug 7, 2025].

21. Benjamin DJ, Berger JO, Johannesson M, Nosek BA, Wagenmakers E-J, Berk R, et al. Redefine statistical significance. Nat Hum Behav. 2018 Jan;2(1):6–10.

22. Stehlik P, Withers C, Bourke RC, Barnett AG, Brandenburg C, Noble C, et al. Mandatory research projects during medical specialist training in Australia and New Zealand: a survey of trainees’ experiences and reports. Med J Aust. 2025 Mar 17;222(5):231–239

23. Tomlinson OW. Predatory publishing in medical education: a rapid scoping review. BMC Med Educ. 2024 Jan 5;24:33.

