## Supplementary material for "Student Scholarly Research Programs in US Medical Schools: Cross-sectional Web Audit": Multimedia Appendix 1

**Multimedia Appendix 1. Reporting completeness (missingness) of program characteristics**

Values are missing counts per variable (documentation not available on webpages) shown as n/N (%).

| **Variable** | **Overall missing, n/N (%)** | **R1 missing, n/N (%)** | **Non-R1 missing, n/N (%)** | **P (R1 vs non-R1)** | **QS Top-50 missing, n/N (%)** | **Non-Top-50 missing, n/N (%)** | **P (Top-50 vs others)** | **MD missing, n/N (%)** | **DO missing, n/N (%)** | **P (MD vs DO)** |
| --- | --- | --- | --- | --- | --- | --- | --- | --- | --- | --- |
| Any funding mentioned | 77/202 (38.1%) | 40/104 (38.5%) | 37/98 (37.8%) | >.999 | 11/19 (57.9%) | 66/183 (36.1%) | .106 | 61/158 (38.6%) | 16/44 (36.4%) | .924 |
| Compulsory participation | 13/202 (6.4%) | 3/104 (2.9%) | 10/98 (10.2%) | **.044** | 0/19 (0.0%) | 13/183 (7.1%) | .616 | 7/158 (4.4%) | 6/44 (13.6%) | .064 |
| Contains summer project | 13/202 (6.4%) | 3/104 (2.9%) | 10/98 (10.2%) | **.044** | 0/19 (0.0%) | 13/183 (7.1%) | .616 | 7/158 (4.4%) | 6/44 (13.6%) | .064 |
| Mentored by faculty | 12/202 (5.9%) | 4/104 (3.8%) | 8/98 (8.2%) | .241 | 0/19 (0.0%) | 12/183 (6.6%) | .609 | 8/158 (5.1%) | 4/44 (9.1%) | .298 |
| Exclusive for medical students | 14/202 (6.9%) | 3/104 (2.9%) | 11/98 (11.2%) | **.026** | 0/19 (0.0%) | 14/183 (7.7%) | .371 | 7/158 (4.4%) | 7/44 (15.9%) | .021 |
| Program director mentioned | 101/202 (50.0%) | 46/104 (44.2%) | 55/98 (56.1%) | .121 | 10/19 (52.6%) | 91/183 (49.7%) | >.999 | 73/158 (46.2%) | 28/44 (63.6%) | .061 |
| Starting year | 47/202 (23.3%) | 14/104 (13.5%) | 33/98 (33.7%) | **.001** | 1/19 (5.3%) | 46/183 (25.1%) | .082 | 24/158 (15.2%) | 23/44 (52.3%) | **P<.001** |
| Ending year | 52/202 (25.7%) | 18/104 (17.3%) | 34/98 (34.7%) | **.008** | 3/19 (15.8%) | 49/183 (26.8%) | .412 | 29/158 (18.4%) | 23/44 (52.3%) | **P<.001** |
| Duration | 30/202 (14.9%) | 9/104 (8.7%) | 21/98 (21.4%) | **.019** | 2/19 (10.5%) | 28/183 (15.3%) | .745 | 16/158 (10.1%) | 14/44 (31.8%) | **P<.001** |
| Dedicated duration | 77/202 (38.1%) | 33/104 (31.7%) | 44/98 (44.9%) | **.075** | 3/19 (15.8%) | 74/183 (40.4%) | **.046** | 54/158 (34.2%) | 23/44 (52.3%) | **.044** |
| Form of final outcome | 37/202 (18.3%) | 15/104 (14.4%) | 22/98 (22.4%) | .196 | 3/19 (15.8%) | 34/183 (18.6%) | >.999 | 29/158 (18.4%) | 8/44 (18.2%) | >.999 |
| Amount of stipend | 141/202 (69.8%) | 68/104 (65.4%) | 73/98 (74.5%) | .209 | 18/19 (94.7%) | 123/183 (67.2%) | **.016** | 106/158 (67.1%) | 35/44 (79.5%) | .160 |
| Goal | 33/202 (16.3%) | 15/104 (14.4%) | 18/98 (18.4%) | .570 | 2/19 (10.5%) | 31/183 (16.9%) | .745 | 24/158 (15.2%) | 9/44 (20.5%) | .545 |
