## Supplementary material for "Student Scholarly Research Programs in US Medical Schools: Cross-sectional Web Audit": Multimedia Appendix 2

### Multimedia Appendix 2. STROBE checklist for cross-sectional studies

Checklist items are based on the STROBE Statement for cross-sectional studies. Please populate the final column with the page/section where each item is addressed in the manuscript.

| Section | Item | Recommendation | Location in manuscript (page/section) |
|---|---|---|---|
| Title and abstract | 1a | Indicate the study's design with a commonly used term in the title or the abstract. | Title; Abstract (first sentence includes study design) |
| Title and abstract | 1b | Provide in the abstract an informative and balanced summary of what was done and what was found. | Abstract (structured: Background/Objective/Methods/Results/Conclusions) |
| Introduction | 2 | Explain the scientific background and rationale for the investigation being reported. | Introduction (background and rationale) |
| Introduction | 3 | State specific objectives, including any prespecified hypotheses. | Introduction: Objective |
| Methods | 4 | Present key elements of study design early in the paper. | Methods: Study Design |
| Methods | 5 | Describe the setting, locations, and relevant dates, including periods of recruitment, exposure, follow-up, and data collection. | Methods: Study Design (setting/dates); Methods: Search Strategy and Data Extraction (search procedures) |
| Methods | 6a | Give the eligibility criteria, and the sources and methods of selection of participants. | Methods: Search Strategy and Data Extraction (institution list; webpage identification) |
| Methods | 6b | For matched studies, give matching criteria and number of exposed and unexposed. | NA (not a matched study) |
| Methods | 7 | Clearly define all outcomes, exposures, predictors, potential confounders, and effect modifiers. Give diagnostic criteria, if applicable. | Methods: Data Extraction (variables/outcomes defined) |
| Methods | 8 | For each variable of interest, give sources of data and details of methods of assessment (measurement). Describe comparability of assessment methods if there is more than one group. | Methods: Search Strategy and Data Extraction; Methods: Data Extraction (webpage sources and coding approach) |

| | | | |
|---|---|---|---|
| Methods | 9 | Describe any efforts to address potential sources of bias. | Methods: Search Strategy and Data Extraction (standardized approach); Discussion: Limitations (misclassification risk) |
| Methods | 10 | Explain how the study size was arrived at. | Methods: Study Design (census of eligible institutions) |
| Methods | 11 | Explain how quantitative variables were handled in the analyses. If applicable, describe which groupings were chosen and why. | Methods: Statistical Analysis |
| Methods | 12a | Describe all statistical methods, including those used to control for confounding. | Methods: Statistical Analysis |
| Methods | 12b | Describe any methods used to examine subgroups and interactions. | Methods: Statistical Analysis (subgroup comparisons by strata) |
| Methods | 12c | Explain how missing data were addressed. | Methods: Data Extraction (items coded as not reported when absent on webpages) |
| Methods | 12d | If applicable, describe analytical methods taking account of sampling strategy. | NA (no sampling strategy) |
| Methods | 12e | Describe any sensitivity analyses. | Not reported (no sensitivity analyses described) |
| Results | 13a | Report numbers of individuals at each stage of study (eg, numbers potentially eligible, examined for eligibility, confirmed eligible, included in the study, completing follow-up, and analysed). | Results: Webpage identification/coverage |
| Results | 13b | Give reasons for non-participation at each stage. | NA |
| Results | 13c | Consider use of a flow diagram. | Not included (optional) |
| Results | 14a | Give characteristics of study participants (eg, demographic, clinical, social) and information on exposures and potential confounders. | Results: Program characteristics: overall; Program characteristics in institutional subgroups; Table 1; Multimedia Appendix 1 |
| Results | 14b | Indicate number of participants with missing data for each variable of interest. | Results: Webpage identification/coverage; Multimedia Appendix 1 |

| | | | |
|---|---|---|---|
| Results | 15 | Report numbers of outcome events or summary measures. | Results: Program characteristics: overall; Program characteristics in institutional subgroups; Tables 1-3 |
| Results | 16a | Give unadjusted estimates and, if applicable, confounder-adjusted estimates and their precision (eg, 95% CI). Make clear which confounders were adjusted for and why they were included. | Results: Program characteristics in institutional subgroups (effect sizes); Tables 1-3 |
| Results | 16b | Report category boundaries when continuous variables were categorized. | Methods: Statistical Analysis (definitions of strata/cutoffs and comparisons) |
| Results | 16c | If relevant, consider translating estimates of relative risk into absolute risk for a meaningful time period. | NA |
| Results | 17 | Report other analyses done (eg, analyses of subgroups and interactions, and sensitivity analyses). | Results: Program characteristics in institutional subgroups (subgroup analyses); Tables 1-3 |
| Discussion | 18 | Summarise key results with reference to study objectives. | Discussion: Principal Findings and Institutional Differences; Conclusions |
| Discussion | 19 | Discuss limitations of the study, taking into account sources of potential bias or imprecision. Discuss both direction and magnitude of any potential bias. | Discussion: Limitations |
| Discussion | 20 | Give a cautious overall interpretation of results considering objectives, limitations, multiplicity of analyses, results from similar studies, and other relevant evidence. | Discussion: Principal Findings and Institutional Differences; Future Directions |
| Discussion | 21 | Discuss the generalisability (external validity) of the study results. | Discussion: Future Directions (external validity/generalizability considerations) |
| Other information | 22 | Give the source of funding and the role of the funders for the present study and, if applicable, for the original study on which the present article is based. | Funding (source of funding and roles) |
